# Effects of System-Level Factors on Race/Ethnic Differences in In-Hospital Mortality after Acute Ischemic Stroke

**DOI:** 10.1101/2023.10.20.23297343

**Authors:** Philip Sun, Daniela Markovic, Abdullah Ibish, Roland Faigle, Rebecca Gottesman, Amytis Towfighi

**Affiliations:** Department of Neurology, David Geffen School of Medicine at University of California – Los Angeles, Los Angeles, CA; Keck School of Medicine of University of Southern California, Los Angeles, CA; Department of Internal Medicine, University of California - Los Angeles, Los Angeles, CA; Department of Neurology, Johns Hopkins University School of Medicine, Baltimore, MD

**Keywords:** thrombolysis, sex differences, ethnicity, elderly, epidemiology, Stroke, Mortality, Disparities, Race, Ethnicity, Inequities

## Abstract

**Introduction:** Stroke mortality has declined, with differential changes by race; stroke is now the 5th leading cause of death overall, but 2nd leading cause of death in Black individuals. Little is known about recent race/ethnic and sex trends in in-hospital mortality after acute ischemic stroke (AIS) and whether system-level factors contribute to possible differences.

**Methods:** Using the National Inpatient Sample, adults (≥18 years) with a primary diagnosis of AIS from 2006 to 2017 (n=643,912) were identified. We assessed in-hospital mortality by race/ethnicity (White, Black, Hispanic, Asian/Pacific Islander [API], other), sex, and age. Hospitals were categorized by proportion of non-White patients served: <25% (“predominantly White patients”), 25-50% (“mixed race/ethnicity profile”), and ≥50% (“predominantly non-White patients”). Using survey adjusted logistic regression, the association between race/ethnicity and odds of mortality was assessed, adjusting for key sociodemographic, clinical, and hospital characteristics (e.g., age, comorbidities, stroke severity, do not resuscitate orders, and palliative care).

**Results:** Overall, mortality decreased from 5.0% in 2006 to 2.9% in 2017 (p<0.001). Comparing 2012-2017 to 2006-2011, there was a 68% reduction in mortality odds overall after adjusting for covariates, most prominent in White individuals (69%) and smallest in Black individuals (57%). Compared to White patients, Black and Hispanic patients had lower odds of mortality (adjusted odds ratio (aOR) 0.82, 95% CI 0.78-0.87 and aOR 0.93, 95% CI 0.87-1.00), primarily driven by those >65 years (age x ethnicity interaction p < 0.0001). Compared to White men, Black, Hispanic, and API men, and Black women had lower aOR of mortality. The differences in mortality between White and non-White patients were most pronounced in hospitals predominantly serving White patients (aOR 0.80, 0.74-0.87) compared to mixed hospitals (aOR 0.85, 0.79-0.91) and predominantly non-White hospitals (aOR 0.88, 0.81-0.95; interaction effect: p=0.005).

**Discussion:** AIS mortality decreased dramatically in recent years in all race/ethnic subgroups. Overall, non-White AIS patients had lower mortality than their White counterparts, a difference that was most striking in hospitals predominantly serving White patients. Further study is needed to understand these differences and to what extent biological, sociocultural, and system-level factors play a role.

**Category:** Health Services, Quality Improvement, and Patient-Centered Outcomes

## Introduction

Stroke is the 5^th^ leading cause of death overall^1^ but the second leading cause of death in women and Black people in the United States^2^. Stroke mortality has declined in the past decades^2,3^, likely due to improved prevention measures resulting in lower incidence and enhanced acute management resulting in lower case fatality. Yet, mortality reductions have been less pronounced in Black individuals^2,4,5^ and women^2,6^. Black individuals face numerous barriers to care, including difficulty receiving high-quality referral and treatment,^7,8^ treatment by physicians with suboptimal clinical qualifications,^7,9,10^ delays in receiving time-sensitive treatments,^10,11^ and being cared for at institutions with poorer outcomes.^12–14^

It is unclear whether differences in health outcomes by race/ethnicity are due to poorer quality care provided to non-White patients compared with White patients or if non-White patients are more likely to be treated by a physician or hospital where *all* patients receive lower quality care, regardless of race/ethnicity^15,16^. One strategy to answer this question is to look at outcomes by race/ethnicity in hospitals categorized by the proportion of non-White individuals admitted. This strategy was used to explain race/ethnic disparities in intravenous thrombolysis after acute ischemic stroke (AIS)^17^. The study found that non-White men and White women were less likely to receive intravenous thrombolysis if they presented to hospitals predominantly serving non-White individuals compared to hospitals serving predominantly White individuals. Little is known about race/ethnic temporal trends in in-hospital mortality after stroke and the extent to which differences are attributable to system-level factors.

The first aim of this study was to evaluate recent temporal trends in in-hospital mortality after AIS by race/ethnicity between 2006 and 2017 using data from the National Inpatient Sample (NIS). Secondly, we aimed to evaluate system-level contributions to differences in in-hospital mortality by comparing outcomes among hospitals by proportion of non-White patients served. We hypothesized an overall decrease in in-hospital mortality over time and higher mortality in non-White patients, particularly in hospitals predominantly serving non-White patients.

## Methods

### Population for Study

Data were obtained from NIS, developed as part of the Healthcare Cost and Utilization Project^18^. Prior to 2012, the survey was designed to approximate a stratified 20% sample of all United States community hospitals (non-federal, short-term, general, and specialty hospitals) serving adults in the United States. From 2012, the sampling strategy transitioned to 20% of patient discharges from all United States community hospitals excluding rehabilitation and long-term acute care hospitals. All selected hospitals have state inpatient databases with defined strata based on ownership, bed size, teaching status, urban/rural location, and region. Admissions to hospitals with fewer than fifty annual ischemic stroke cases were excluded. All discharges from sampled hospitals for the calendar year are then selected for inclusion into NIS. To allow extrapolation for national estimates, both hospital and discharge weights are provided. Detailed information on the design of the NIS is available at http://www.hcup-us.ahrq.gov.

NIS captures discharge-level information on primary and secondary diagnoses and procedures and demographics on several million discharges per year. Data elements that could directly or indirectly identify individuals are excluded. The unit of analysis was the discharge rather than the individual; discharges were therefore all considered independent. A unique hospital identifier allows for linkage of discharge data to an NIS data set with hospital characteristics.

All hospitalizations among adult patients admitted with a primary or secondary diagnosis AIS (International Classification of Diseases, Ninth Revision diagnosis codes [ICD-9-CM] 433.01, 433.11, 433.21, 433.31, 433.81, 433.91, 434.01, 434.11, 434.91, 436, 437.0, 437.1) and Tenth Revision diagnosis codes [ICD-10-CM] I63.019, I63.119, I63.139, I63.219, I63.20, I63.22, I63.239, I63.30, I63.40, I63.50, I63.59, I67.89) at the time of hospital admission were included. ICD-9 codes were used for admissions through September 2015 and ICD-10 codes were used thereafter. Intravenous thrombolysis was determined using ICD-9 procedure code 99.10 and ICD-10 procedure code 3E03317.

We excluded hospitalizations for patients who were younger than 18 years of age, missing race/ethnicity or sex, had a diagnosis of acute myocardial infarction and/or pulmonary embolism, malignancy (solid tumor without metastasis, lymphoma, metastatic cancer), transferred to index hospital from another hospital, elective admissions, or enrolled in a clinical trial (ICD-9-CM code V70.7, ICD-10-CM code Z006). Please refer to **Supplemental Tables 1-3** for full ICD-9/10 code lists for exclusion criteria and covariates ^19^.

### Hospital Categorization/Outcome measures

Hospitals were categorized by proportion of non-White patients served: <25% (“predominantly White patients”), 25-50% (“mixed racial/ethnic profile”), and ≥50% (“predominantly non-White patients”)^14,17^. Non-White race/ethnicity included all patients identified as Black, Hispanic, Asian/Pacific Islander, or other. The primary outcome measure of interest was race/ethnic-specific in-hospital mortality by hospital categories among patients admitted for AIS. The outcome’s association with age and sex were assessed.

### Sociodemographic, Clinical, and Hospital Factors

Individuals were categorized by age groups: minors (<18 years), young adults (18-44 years), mid-life (45-64 years), elderly (65-84 years), and very elderly (≥85 years). The following factors were assessed: sex (women/men), race/ethnicity (White, Black, Hispanic, Asian/Pacific Islander [API], other), primary payer (Medicare, Medicaid, private including Health Maintenance Organization [HMO], others), hospital location/teaching status (urban teaching, urban non-teaching, rural), hospital region (Northeast, Midwest, South, West), hospital ownership (government nonfederal, private non-profit, private invest-own, others/missing), and hospital bed size (small [≤200 beds], medium [200-400 beds], large [>400 beds]). The following comorbidities were included as covariates: hypertension, dyslipidemia, alcohol abuse, obesity, smoking history, coronary artery disease, atrial fibrillation, and Charlson Comorbidity Index^20^ (consisting of 17 comorbidities; **Supplemental Table 3**). Charlson Comorbidity Index has been validated for outcome adjustment for analysis of administrative data sets using ICD-9 codes^20,21^. A subsequent study described corresponding ICD-10 diagnoses for the 17 factors included in the Charlson Comorbidity Index.^19^ Risk of mortality was determined by the Risk of Mortality Subclass Category^22^. Disease severity was assessed by “All Patient Refined Diagnosis Related Groups (APRDRG) Severity, which is a 4-point ordinal scale of mortality risk (minor, moderate, major, and extreme) derived from age, primary and secondary diagnoses, and procedures. The APRDRG algorithm is a validated and reliable indicator of mortality and is commonly used as a severity indicator in studies relating to stroke^23,24^. ICD-9-CM code of V66.7 identifies documented use of palliative care measures^25^, irrespective of the delivery mode (e.g. via a palliative care consultation service or integrated into routine clinical practice by the care team). V66.7 has been shown to accurately identify palliative care services in stroke patients with 81% sensitivity and 97% specificity^26^. Use of ICD-10-CM code Z51.5 has been validated in palliative care patients recently^27^, but its sensitivity and specificity has not been extensively assessed.

### Statistical Analyses

We assessed in-hospital mortality by race/ethnicity, sex, and age. Temporal trends in race/ethnic-specific in-hospital mortality after AIS were assessed from 2006 through 2017. The survey adjusted logistic regression model was used to compare the odds of in-hospital mortality over time in two ways 1) per year and 2) between time intervals (2^nd^ half vs. 1^st^ half of the time frame), adjusting for age, sex, primary payer, hospital region, hospital location/teaching status, hospital ownership, hospital bed size, APRDRG Severity, Charlson Comorbidity Index, do not resuscitate orders, and palliative treatment.

The survey adjusted logistic regression model was also used to compare the odds of in-hospital mortality across race/ethnicity (White vs. non-White) and sex within each hospital type. Similar methods were used to compare in-hospital mortality across hospital types within each race/ethnicity. Additional subset analyses comparing in-hospital mortality across race/ethnicity were performed for different combinations of age and sex. All incidence estimates were appropriately weighted to reflect U.S. national estimates.

Categorical variables were compared across hospital type using the Rao-Scott Chi-square test, whereas continuous variables were compared across the above groups using linear regression analysis after adjusting for the survey design variables. Analyses were performed using SAS 9.4 (Copyright © 2016 by SAS Institute Inc., Cary, NC, USA). Statistical hypotheses were tested using p<0.05 as the level of statistical significance.

The study was considered exempt from institutional review board given the use of deitentified information. We followed the Strengthening the Reporting of Observational Studies in Epidemiology (STROBE) reporting guideline^28^.

## Results

Among the 643,912 hospitalizations that met inclusion criteria, 328,719 (51.0%) occurred at hospitals predominantly serving White AIS patients, 179,604 (27.9%) at hospitals serving AIS patients with mixed race/ethnicity profiles, and 165,694 (21.7%) at hospitals serving predominantly non-White AIS patients (**Table 1**). The mean age was 71 years, 52% were women, and the mean length of stay was 4.94 +/-0.02 days. **Table 1** shows patient and hospital characteristics by hospital category.

**Table 1.**
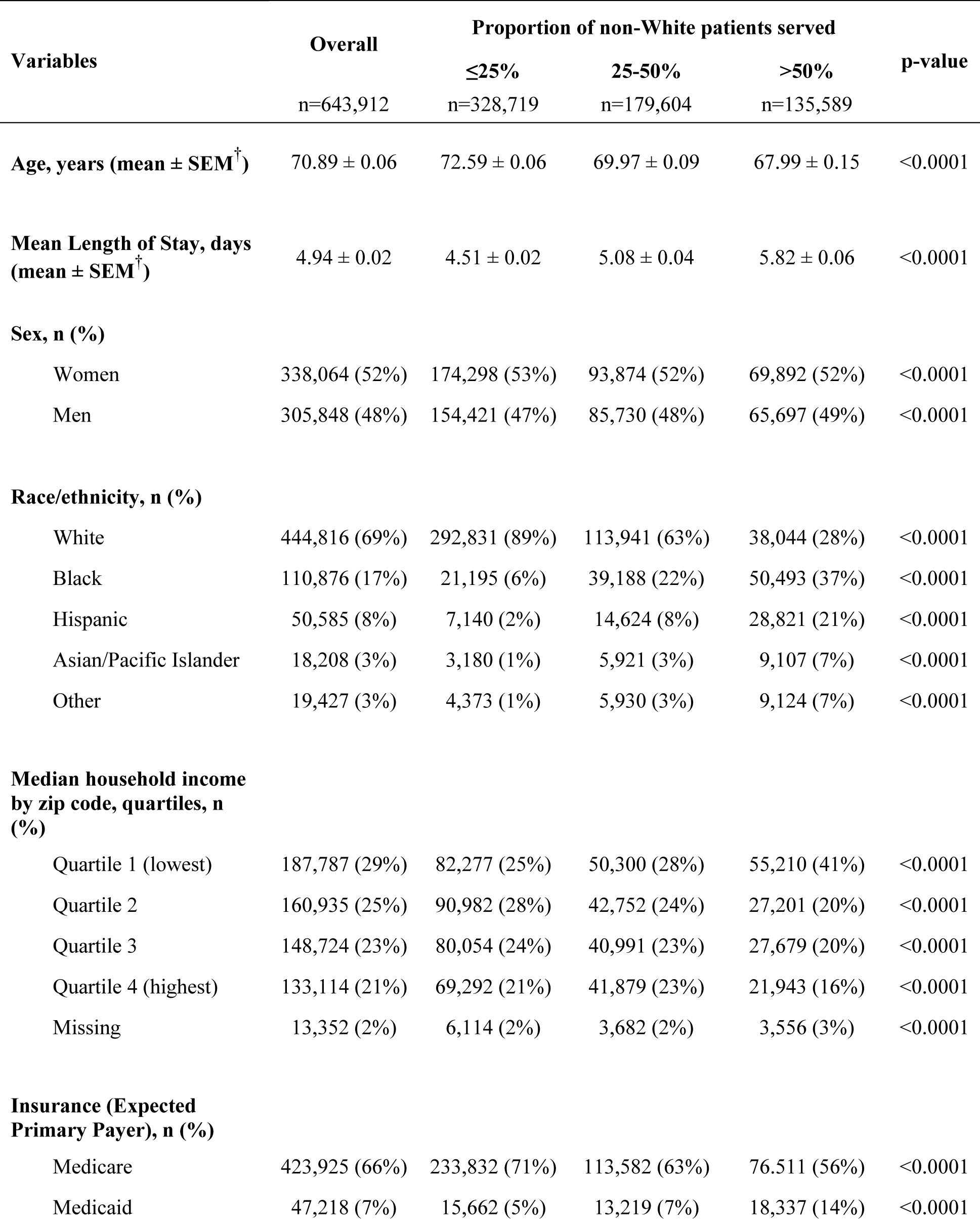
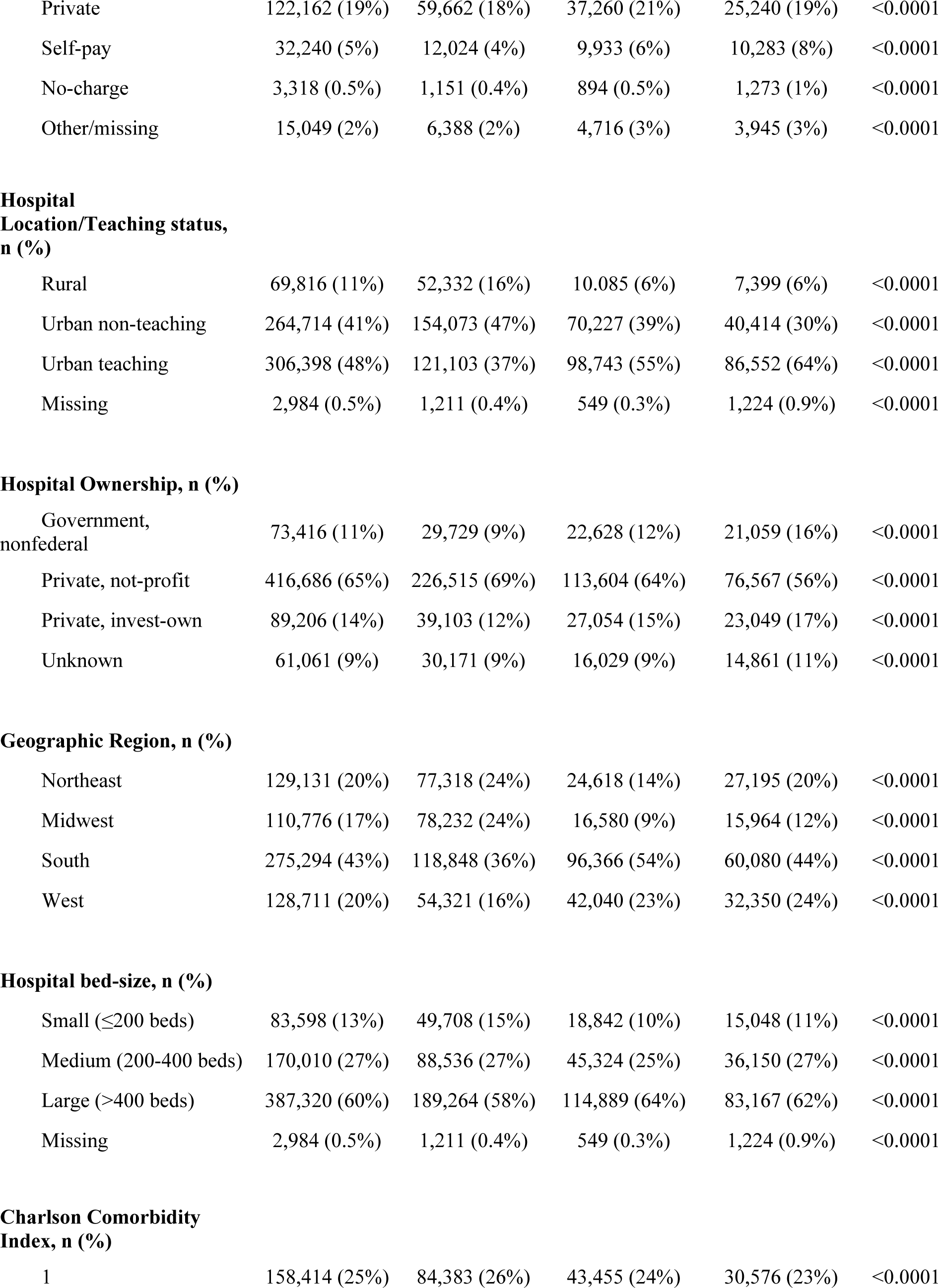
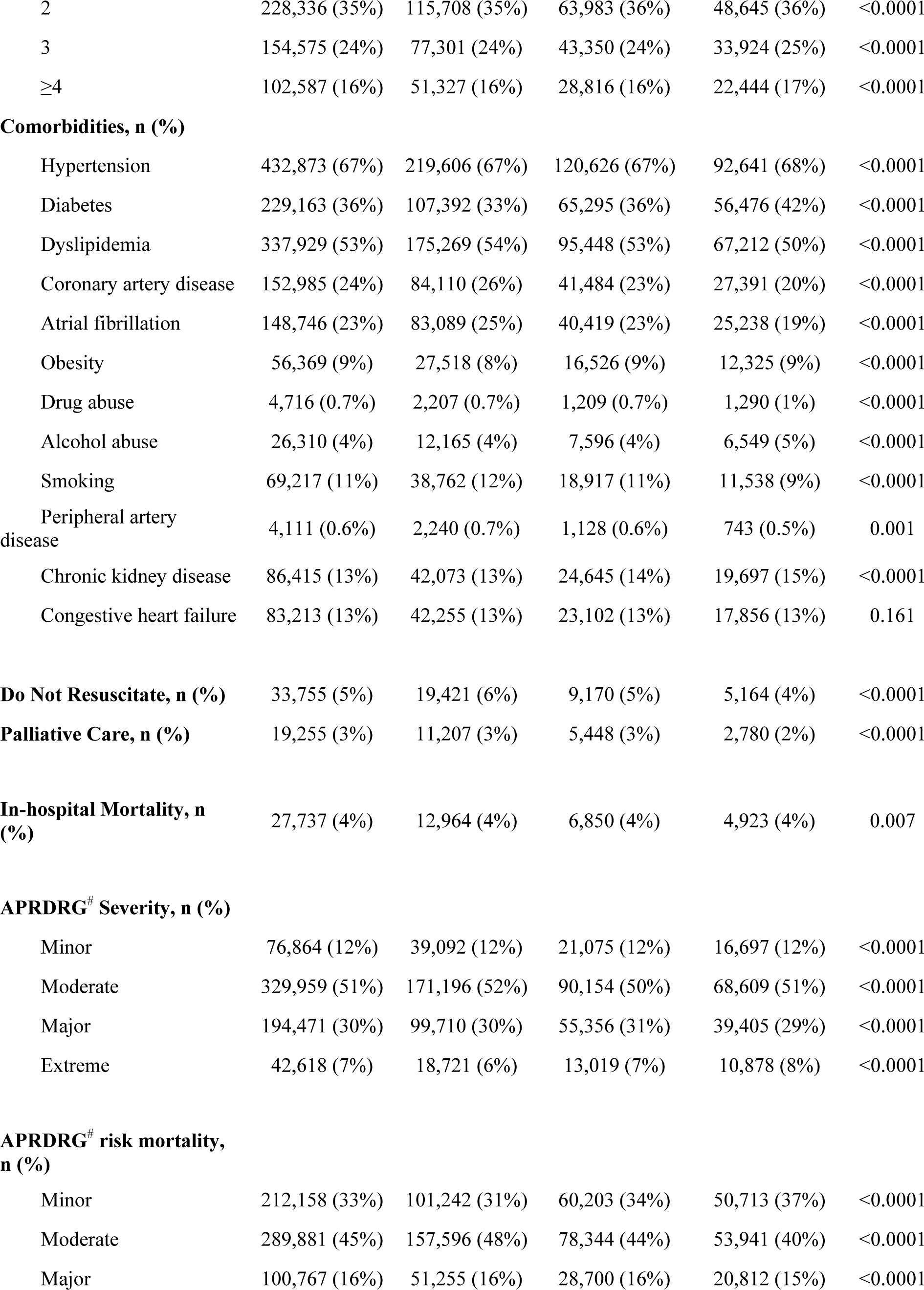
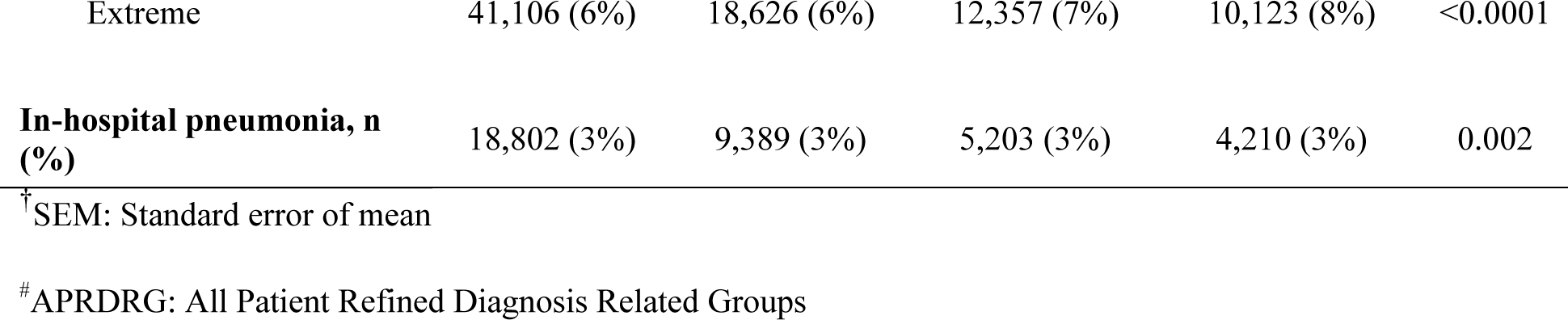
Patient and hospital characteristics for hospitalizations among individuals admitted for acute ischemic stroke, by hospital category, 2006-2017.

| Variables | Overall<br>n=643,912 | Proportion of non-White patients served |  |  | p-value |
| --- | --- | --- | --- | --- | --- |
|  |  | ≤25%<br>n=328,719 | 25-50%<br>n=179,604 | >50%<br>n=135,589 |  |
| **Age, years (mean ± SEM<sup>†</sup>)** | 70.89 ± 0.06 | 72.59 ± 0.06 | 69.97 ± 0.09 | 67.99 ± 0.15 | <0.0001 |
| **Mean Length of Stay, days (mean ± SEM<sup>†</sup>)** | 4.94 ± 0.02 | 4.51 ± 0.02 | 5.08 ± 0.04 | 5.82 ± 0.06 | <0.0001 |
| **Sex, n (%)** |  |  |  |  |  |
| Women | 338,064 (52%) | 174,298 (53%) | 93,874 (52%) | 69,892 (52%) | <0.0001 |
| Men | 305,848 (48%) | 154,421 (47%) | 85,730 (48%) | 65,697 (49%) | <0.0001 |
| **Race/ethnicity, n (%)** |  |  |  |  |  |
| White | 444,816 (69%) | 292,831 (89%) | 113,941 (63%) | 38,044 (28%) | <0.0001 |
| Black | 110,876 (17%) | 21,195 (6%) | 39,188 (22%) | 50,493 (37%) | <0.0001 |
| Hispanic | 50,585 (8%) | 7,140 (2%) | 14,624 (8%) | 28,821 (21%) | <0.0001 |
| Asian/Pacific Islander | 18,208 (3%) | 3,180 (1%) | 5,921 (3%) | 9,107 (7%) | <0.0001 |
| Other | 19,427 (3%) | 4,373 (1%) | 5,930 (3%) | 9,124 (7%) | <0.0001 |
| **Median household income by zip code, quartiles, n (%)** |  |  |  |  |  |
| Quartile 1 (lowest) | 187,787 (29%) | 82,277 (25%) | 50,300 (28%) | 55,210 (41%) | <0.0001 |
| Quartile 2 | 160,935 (25%) | 90,982 (28%) | 42,752 (24%) | 27,201 (20%) | <0.0001 |
| Quartile 3 | 148,724 (23%) | 80,054 (24%) | 40,991 (23%) | 27,679 (20%) | <0.0001 |
| Quartile 4 (highest) | 133,114 (21%) | 69,292 (21%) | 41,879 (23%) | 21,943 (16%) | <0.0001 |
| Missing | 13,352 (2%) | 6,114 (2%) | 3,682 (2%) | 3,556 (3%) | <0.0001 |
| **Insurance (Expected Primary Payer), n (%)** |  |  |  |  |  |
| Medicare | 423,925 (66%) | 233,832 (71%) | 113,582 (63%) | 76,511 (56%) | <0.0001 |
| Medicaid | 47,218 (7%) | 15,662 (5%) | 13,219 (7%) | 18,337 (14%) | <0.0001 |
| Private | 122,162 (19%) | 59,662 (18%) | 37,260 (21%) | 25,240 (19%) | <0.0001 |
| Self-pay | 32,240 (5%) | 12,024 (4%) | 9,933 (6%) | 10,283 (8%) | <0.0001 |
| No-charge | 3,318 (0.5%) | 1,151 (0.4%) | 894 (0.5%) | 1,273 (1%) | <0.0001 |
| Other/missing | 15,049 (2%) | 6,388 (2%) | 4,716 (3%) | 3,945 (3%) | <0.0001 |
| **Hospital Location/Teaching status, n (%)** |  |  |  |  |  |
| Rural | 69,816 (11%) | 52,332 (16%) | 10,085 (6%) | 7,399 (6%) | <0.0001 |
| Urban non-teaching | 264,714 (41%) | 154,073 (47%) | 70,227 (39%) | 40,414 (30%) | <0.0001 |
| Urban teaching | 306,398 (48%) | 121,103 (37%) | 98,743 (55%) | 86,552 (64%) | <0.0001 |
| Missing | 2,984 (0.5%) | 1,211 (0.4%) | 549 (0.3%) | 1,224 (0.9%) | <0.0001 |
| **Hospital Ownership, n (%)** |  |  |  |  |  |
| Government, nonfederal | 73,416 (11%) | 29,729 (9%) | 22,628 (12%) | 21,059 (16%) | <0.0001 |
| Private, not-profit | 416,686 (65%) | 226,515 (69%) | 113,604 (64%) | 76,567 (56%) | <0.0001 |
| Private, invest-own | 89,206 (14%) | 39,103 (12%) | 27,054 (15%) | 23,049 (17%) | <0.0001 |
| Unknown | 61,061 (9%) | 30,171 (9%) | 16,029 (9%) | 14,861 (11%) | <0.0001 |
| **Geographic Region, n (%)** |  |  |  |  |  |
| Northeast | 129,131 (20%) | 77,318 (24%) | 24,618 (14%) | 27,195 (20%) | <0.0001 |
| Midwest | 110,776 (17%) | 78,232 (24%) | 16,580 (9%) | 15,964 (12%) | <0.0001 |
| South | 275,294 (43%) | 118,848 (36%) | 96,366 (54%) | 60,080 (44%) | <0.0001 |
| West | 128,711 (20%) | 54,321 (16%) | 42,040 (23%) | 32,350 (24%) | <0.0001 |
| **Hospital bed-size, n (%)** |  |  |  |  |  |
| Small ($\leq 200$ beds) | 83,598 (13%) | 49,708 (15%) | 18,842 (10%) | 15,048 (11%) | <0.0001 |
| Medium (200-400 beds) | 170,010 (27%) | 88,536 (27%) | 45,324 (25%) | 36,150 (27%) | <0.0001 |
| Large ($> 400$ beds) | 387,320 (60%) | 189,264 (58%) | 114,889 (64%) | 83,167 (62%) | <0.0001 |
| Missing | 2,984 (0.5%) | 1,211 (0.4%) | 549 (0.3%) | 1,224 (0.9%) | <0.0001 |
| **Charlson Comorbidity Index, n (%)** |  |  |  |  |  |
| 1 | 158,414 (25%) | 84,383 (26%) | 43,455 (24%) | 30,576 (23%) | <0.0001 |
| 2 | 228,336 (35%) | 115,708 (35%) | 63,983 (36%) | 48,645 (36%) | <0.0001 |
| 3 | 154,575 (24%) | 77,301 (24%) | 43,350 (24%) | 33,924 (25%) | <0.0001 |
| ≥4 | 102,587 (16%) | 51,327 (16%) | 28,816 (16%) | 22,444 (17%) | <0.0001 |
| **Comorbidities, n (%)** |  |  |  |  |  |
| Hypertension | 432,873 (67%) | 219,606 (67%) | 120,626 (67%) | 92,641 (68%) | <0.0001 |
| Diabetes | 229,163 (36%) | 107,392 (33%) | 65,295 (36%) | 56,476 (42%) | <0.0001 |
| Dyslipidemia | 337,929 (53%) | 175,269 (54%) | 95,448 (53%) | 67,212 (50%) | <0.0001 |
| Coronary artery disease | 152,985 (24%) | 84,110 (26%) | 41,484 (23%) | 27,391 (20%) | <0.0001 |
| Atrial fibrillation | 148,746 (23%) | 83,089 (25%) | 40,419 (23%) | 25,238 (19%) | <0.0001 |
| Obesity | 56,369 (9%) | 27,518 (8%) | 16,526 (9%) | 12,325 (9%) | <0.0001 |
| Drug abuse | 4,716 (0.7%) | 2,207 (0.7%) | 1,209 (0.7%) | 1,290 (1%) | <0.0001 |
| Alcohol abuse | 26,310 (4%) | 12,165 (4%) | 7,596 (4%) | 6,549 (5%) | <0.0001 |
| Smoking | 69,217 (11%) | 38,762 (12%) | 18,917 (11%) | 11,538 (9%) | <0.0001 |
| Peripheral artery disease | 4,111 (0.6%) | 2,240 (0.7%) | 1,128 (0.6%) | 743 (0.5%) | 0.001 |
| Chronic kidney disease | 86,415 (13%) | 42,073 (13%) | 24,645 (14%) | 19,697 (15%) | <0.0001 |
| Congestive heart failure | 83,213 (13%) | 42,255 (13%) | 23,102 (13%) | 17,856 (13%) | 0.161 |
| **Do Not Resuscitate, n (%)** | 33,755 (5%) | 19,421 (6%) | 9,170 (5%) | 5,164 (4%) | <0.0001 |
| **Palliative Care, n (%)** | 19,255 (3%) | 11,207 (3%) | 5,448 (3%) | 2,780 (2%) | <0.0001 |
| **In-hospital Mortality, n (%)** | 27,737 (4%) | 12,964 (4%) | 6,850 (4%) | 4,923 (4%) | 0.007 |
| **APRDRG<sup>#</sup> Severity, n (%)** |  |  |  |  |  |
| Minor | 76,864 (12%) | 39,092 (12%) | 21,075 (12%) | 16,697 (12%) | <0.0001 |
| Moderate | 329,959 (51%) | 171,196 (52%) | 90,154 (50%) | 68,609 (51%) | <0.0001 |
| Major | 194,471 (30%) | 99,710 (30%) | 55,356 (31%) | 39,405 (29%) | <0.0001 |
| Extreme | 42,618 (7%) | 18,721 (6%) | 13,019 (7%) | 10,878 (8%) | <0.0001 |
| **APRDRG<sup>#</sup> risk mortality, n (%)** |  |  |  |  |  |
| Minor | 212,158 (33%) | 101,242 (31%) | 60,203 (34%) | 50,713 (37%) | <0.0001 |
| Moderate | 289,881 (45%) | 157,596 (48%) | 78,344 (44%) | 53,941 (40%) | <0.0001 |
| Major | 100,767 (16%) | 51,255 (16%) | 28,700 (16%) | 20,812 (15%) | <0.0001 |
| Extreme | 41,106 (6%) | 18,626 (6%) | 12,357 (7%) | 10,123 (8%) | <0.0001 |
| **In-hospital pneumonia, n (%)** | 18,802 (3%) | 9,389 (3%) | 5,203 (3%) | 4,210 (3%) | 0.002 |

<sup>†</sup>SEM: Standard error of mean
<sup>#</sup>APRDRG: All Patient Refined Diagnosis Related Groups

In-hospital mortality rates decreased from 5.0% in 2006 to 2.9% in 2017 (p<0.001). Adjusted annual plots showed a decreasing trend for all race/ethnicity groups in all hospital categories and overall (**Figure 1a-1d**). The changes differed by race/ethnicity, with the annual percent reduction in in-hospital mortality odds ranging from 12.0 to 15.7% across groups (race/ethnicity x year interaction: p=0.015. Comparing 2012-2017 to 2006-2011, there was a 67% reduction in in-hospital mortality odds overall after adjusting for covariates, most prominent in White individuals (69%) and least prominent in Black individuals (57%) (**Table 2**). Use of DNR orders and palliative treatment increased over the study period (**Supplemental Table 4**).

**Figure 1.**
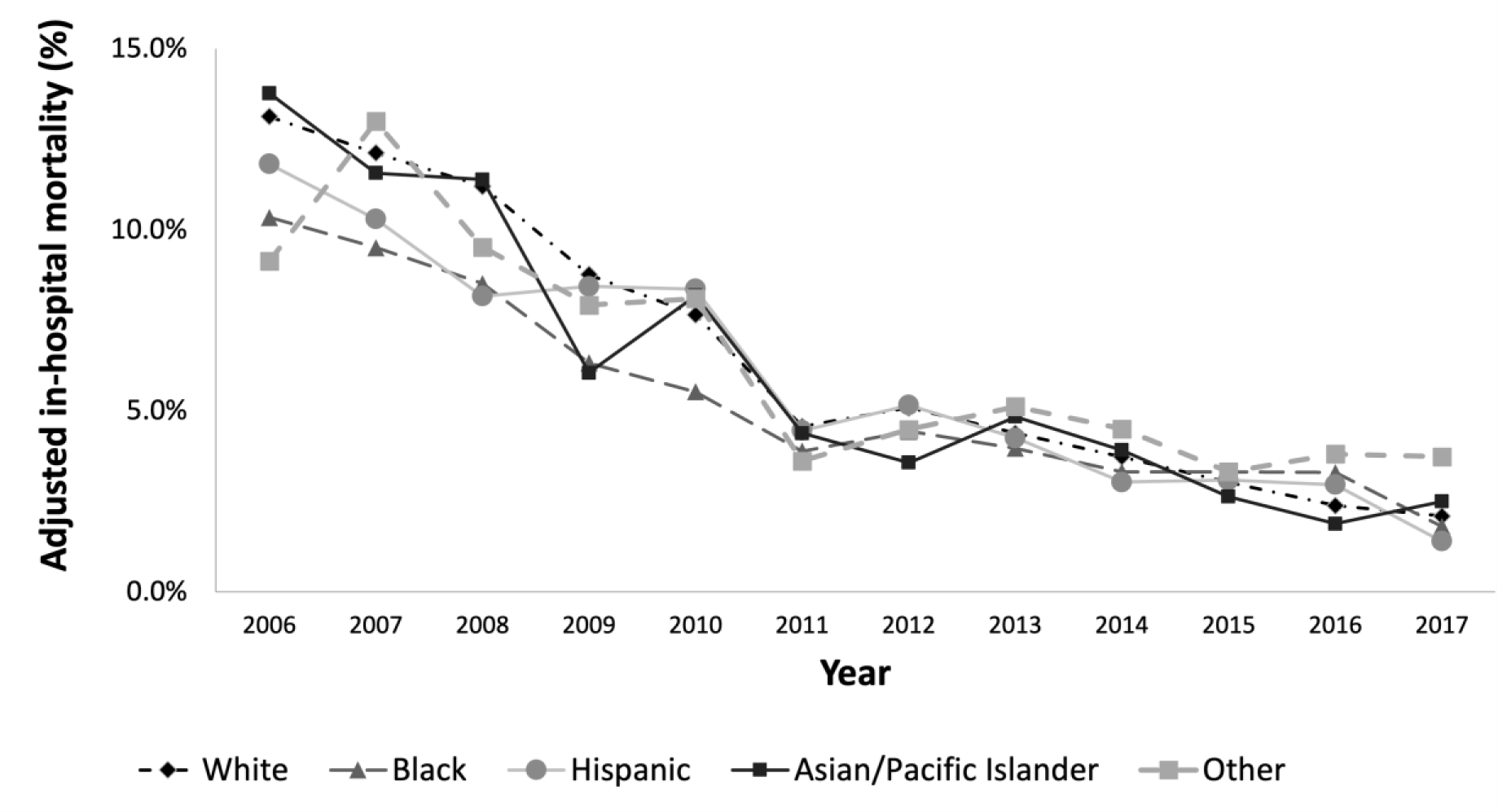
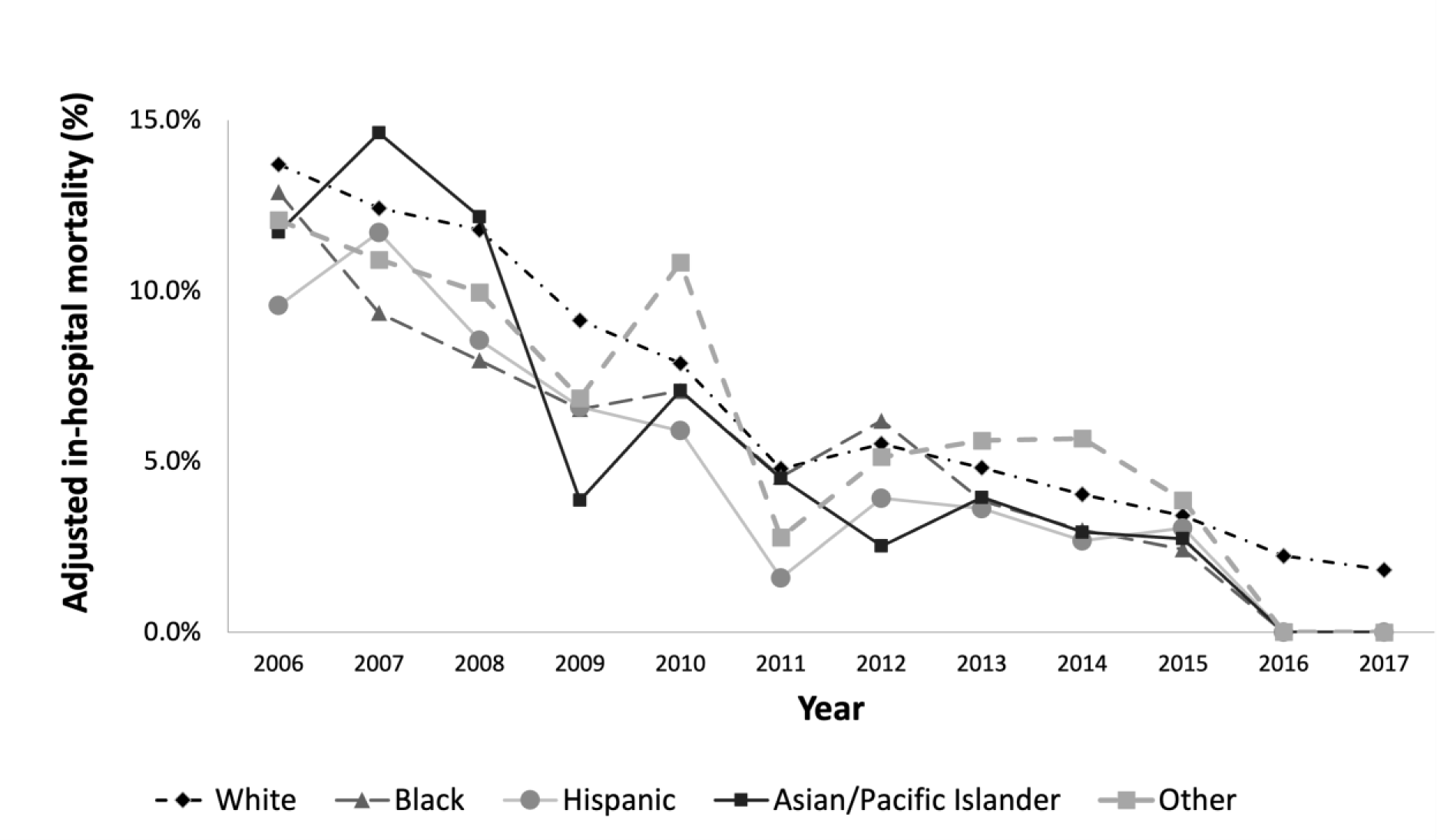
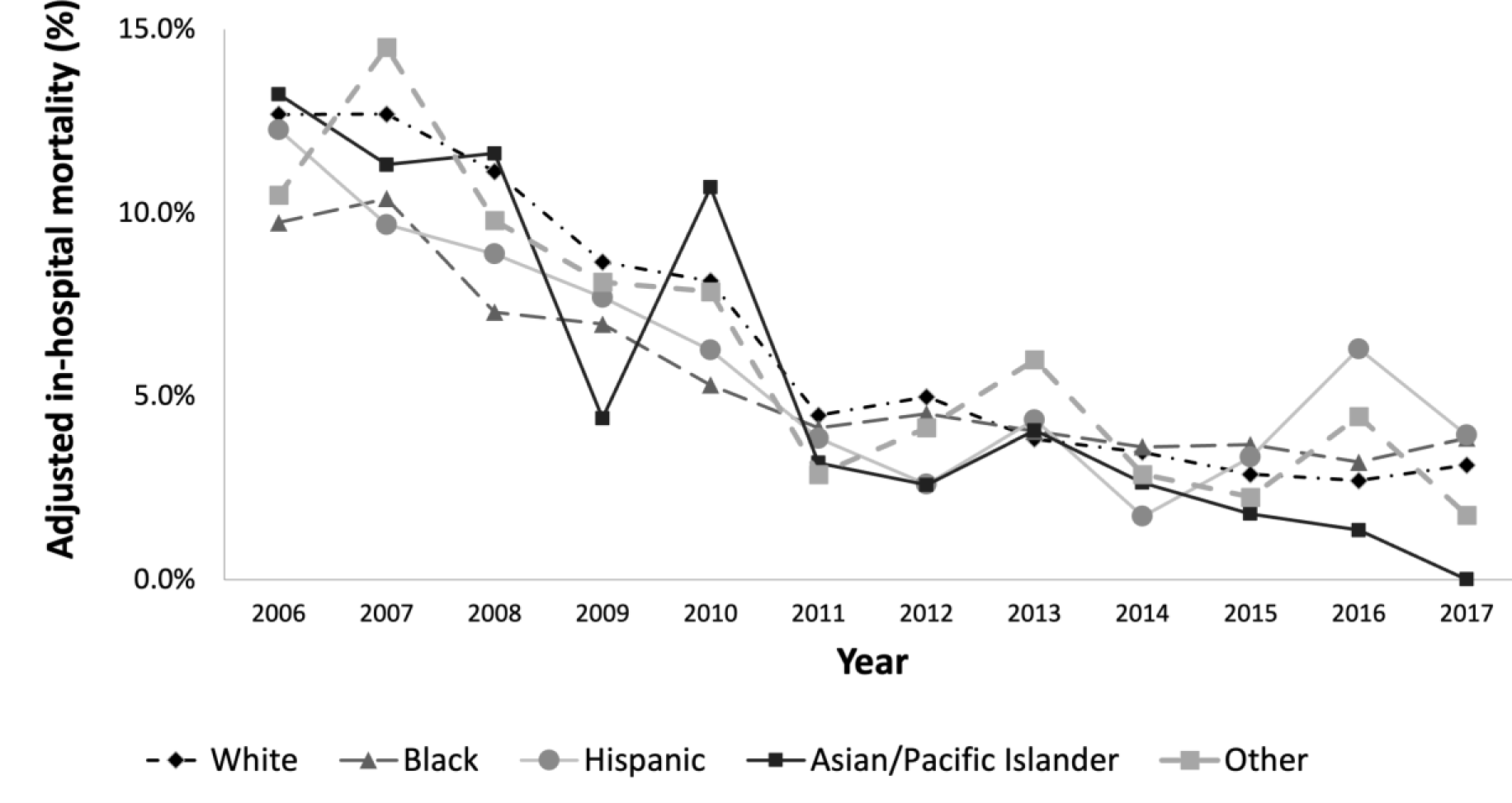
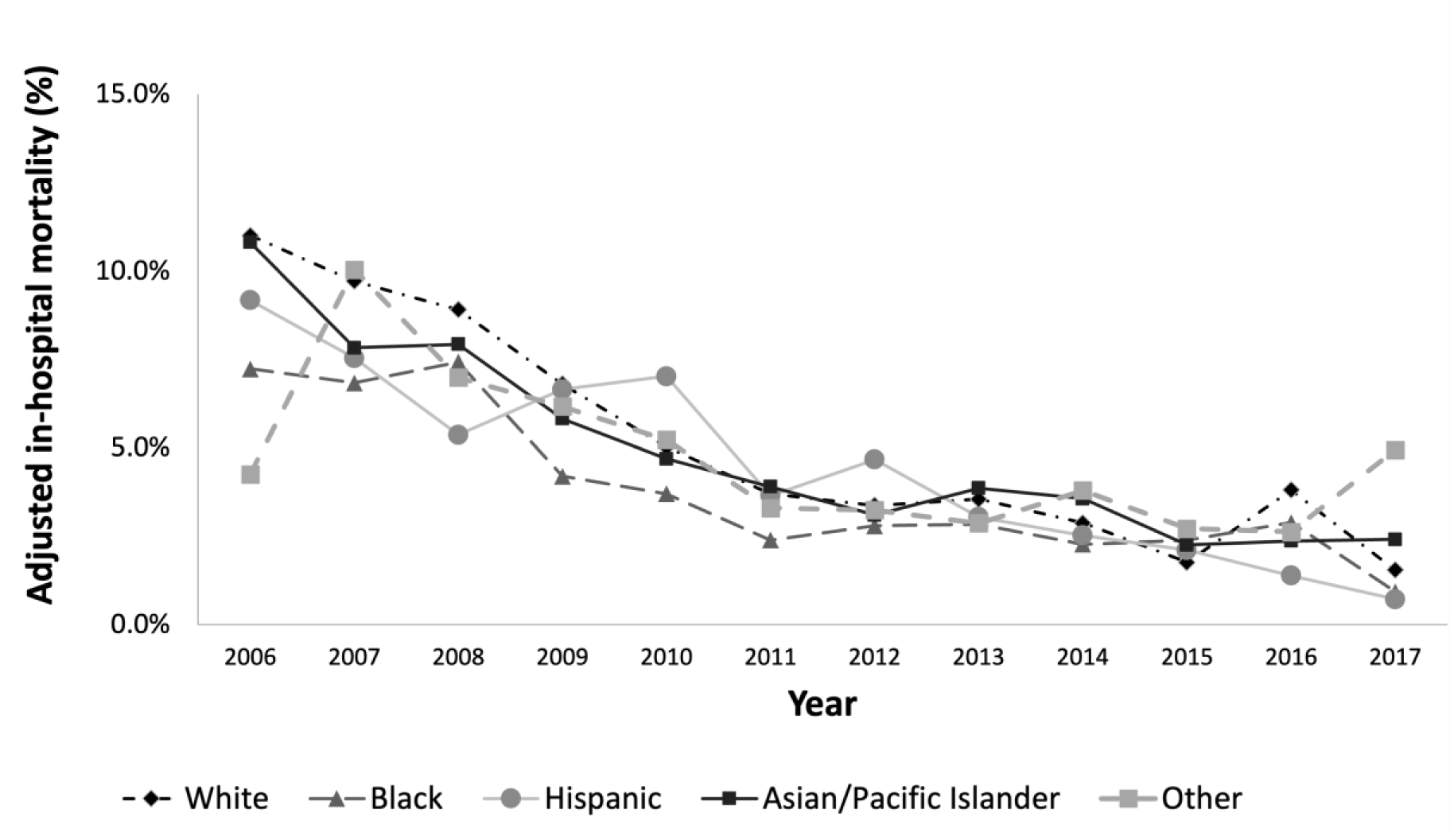
Temporal trends in in-hospital mortality after acute ischemic stroke by race/ethnicity between 2006-2017 in a) all hospitals, b) hospitals serving predominantly White patients, c) hospitals with mixed race/ethnic profile, and d) hospitals serving predominantly non-White patients. All were adjusted for sex, insurance, hospital region, hospital location/teaching status, hospital ownership, bed size, All Patient Refined Diagnosis Related Groups (APRDRG) Severity, Charlson Comorbidity score, do not resuscitate (DNR), and palliative care.

**Table 2.** Adjusted comparisons of in-hospital mortality odds over time by race/ethnicity from 2006 to 2017.

| **Temporal changes in in-hospital mortality** |  |  |  |  |  |  |
| --- | --- | --- | --- | --- | --- | --- |
| **Race/ethnicity** | *2012-2017 vs. 2006-2011* |  |  | Adjusted annual % rate of decline in mortality odds |  |  |
|  | Unadjusted OR† | Adjusted OR† | p-value | Unadjusted % decline (95% CI) | Adjusted % decline (95% CI) | p-value |
| **White** | 0.65<br>(0.61, 0.70) | 0.31<br>(0.28, 0.34) | <0.001 | 5.2<br>(4.6, 5.9) | 15.7<br>(14.8, 16.6) | <0.001 |
| **Black** | 0.72<br>(0.61, 0.84) | 0.43<br>(0.36, 0.52) | <0.001 | 6.2<br>(4.7, 7.6) | 13.1<br>(11.1, 14.9) | <0.001 |
| **Hispanic** | 0.67<br>(0.54, 0.84) | 0.28<br>(0.21, 0.39) | <0.001 | 4.6<br>(3.0, 6.3) | 16.0<br>(13.6, 18.3) | <0.001 |
| **Asian/Pacific Islander** | 0.69<br>(0.49, 0.95) | 0.29<br>(0.19, 0.44) | <0.001 | 5.1<br>(2.3, 7.8) | 15.0<br>(11.3, 18.5) | <0.001 |
| **Other** | 0.87<br>(0.64, 1.19) | 0.43<br>(0.27, 0.70) | 0.001 | 2.1<br>(-0.7, 4.8) | 12.0<br>(8.0, 15.8) | <0.001 |
| **Overall** | 0.67<br>(0.63, 0.72) | 0.32<br>(0.30, 0.35) | <0.001 | 5.3<br>(4.7, 5.9) | 15.3<br>(14.5, 16.1) | <0.001 |

† Odds Ratio was adjusted for age, sex, primary payer, hospital region, hospital location/teaching status, hospital ownership, hospital bed size, APRDRG Severity, Charlson Comorbidity Index, DNR, and palliative treatment.

Compared to White individuals, Black and Hispanic patients had a significantly lower aOR of mortality, (aOR 0.82, 95% CI 0.78-0.87 and aOR 0.93, 95% CI 0.87-1.00, respectively), with a significantly interaction between age and race/ethnicity (p < 0.0001) but not between race/ethnicity and sex (p=0.94). The above associations were driven primarily by older age groups (**Table 3a**). When the patients were assessed by sex and race/ethnicity, Black men and women and API men had lower odds of in-hospital mortality compared to White men (**Table 3b**). This effect was driven by those ≥ 65 years for Black men and women, and by those aged ≥ 85 for API men. For those with race/ethnicity of “Other”, women 65-84 years of age had lower odds of in-hospital mortality, and men ≥85 years of age had higher odds of in-hospital mortality than White men (**Table 3a**).

**Table 3a.**
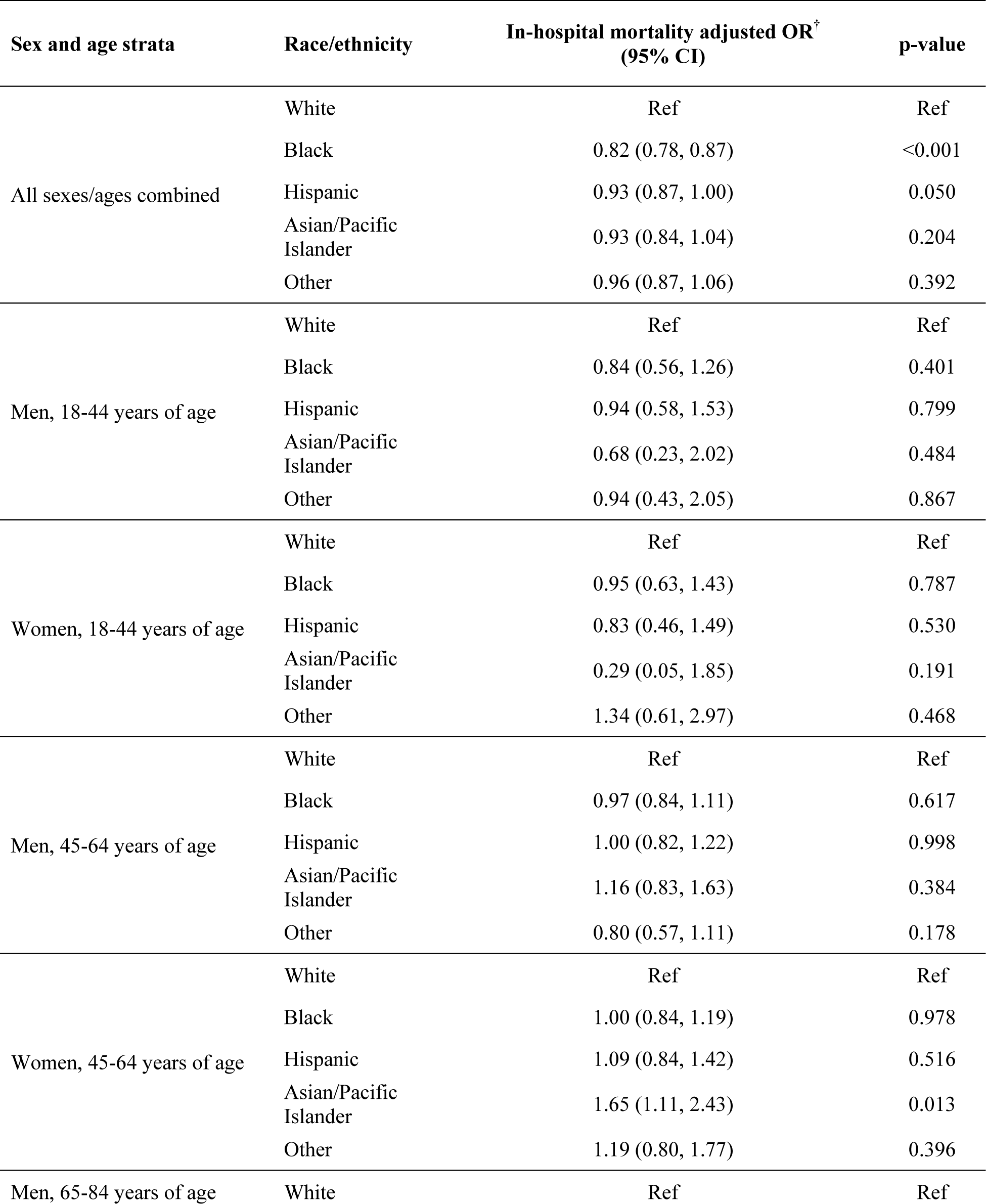
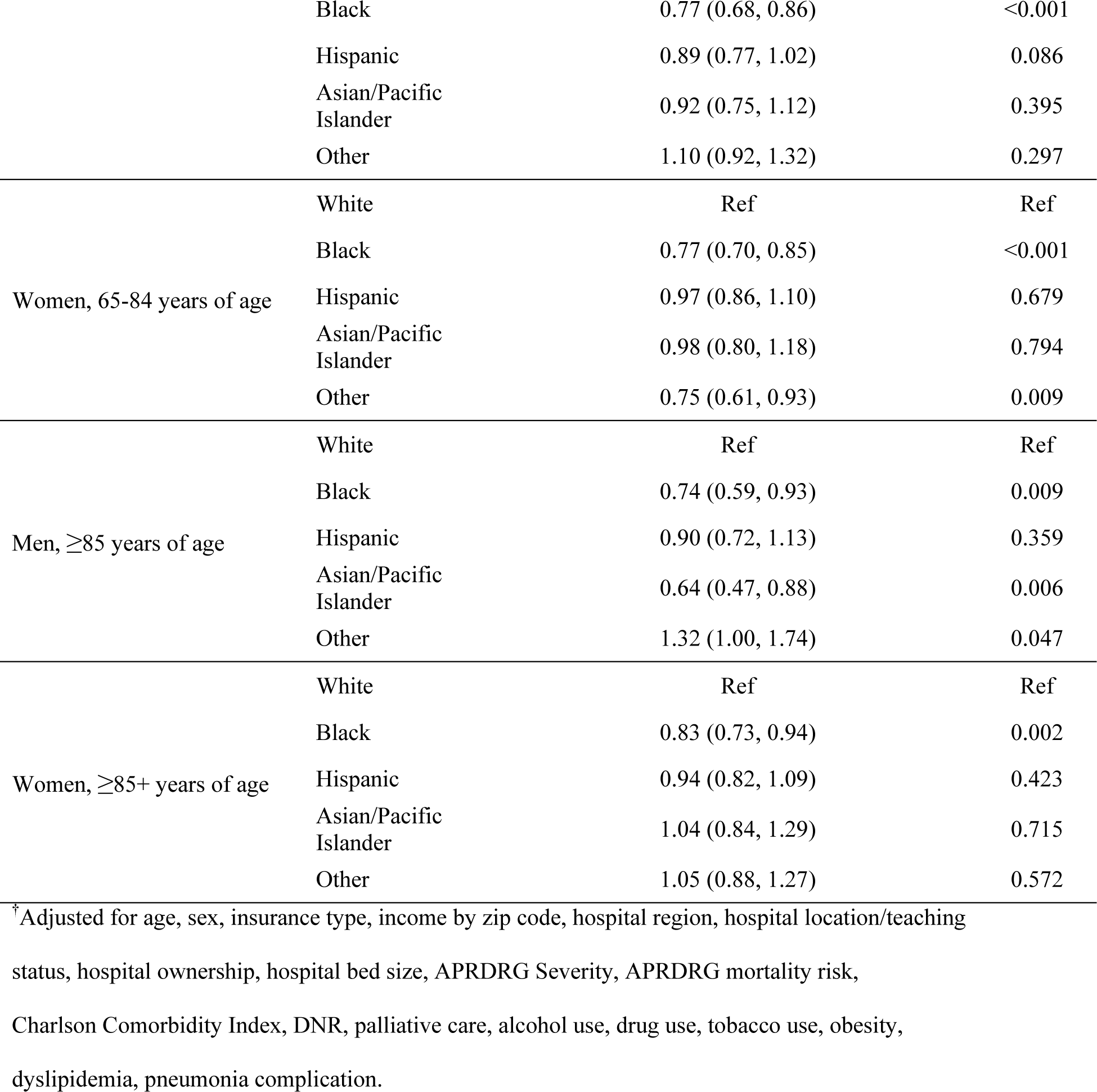
Adjusted comparisons of in-hospital mortality across race/ethnic groups by age and sex strata.

**Table 3b.** Adjusted comparisons of in-hospital mortality across race/ethnicity and sex.

| Race/ethnicity and sex | Odds ratios for in-hospital mortality (95% CI) |  |  |  |
| --- | --- | --- | --- | --- |
|  | unadjusted OR | p-value | adjusted OR<sup>†</sup> | p-value |
| White Men | Ref | Ref | Ref | Ref |
| Black Men | 0.71 (0.67, 0.76) | <0.001 | 0.85 (0.79, 0.91) | <0.001 |
| Hispanic Men | 0.83 (0.77, 0.91) | <0.001 | 0.92 (0.83, 1.01) | 0.088 |
| Asian/Pacific Men | 0.87 (0.77, 0.98) | 0.019 | 0.85 (0.73, 0.98) | 0.024 |
| Other Men | 0.99 (0.88, 1.10) | 0.814 | 1.04 (0.91, 1.18) | 0.618 |
| White Women | 1.25 (1.21, 1.29) | <0.001 | 1.02 (0.98, 1.06) | 0.319 |
| Black Women | 0.76 (0.72, 0.81) | <0.001 | 0.82 (0.77, 0.88) | <0.001 |
| Hispanic Women | 1.04 (0.96, 1.12) | 0.330 | 0.97 (0.88, 1.06) | 0.441 |
| Asian/Pacific Women | 1.29 (1.15, 1.45) | <0.001 | 1.02 (0.88, 1.19) | 0.755 |
| Other Women | 1.15 (1.03, 1.27) | 0.014 | 0.91 (0.79, 1.06) | 0.217 |

<sup>†</sup> Adjusted for age, sex, insurance type, income by zip code, hospital region, hospital location/teaching status, hospital ownership, hospital bed size, APRDRG Severity, APRDRG mortality risk, Charlson Comorbidity Index, DNR, palliative care, alcohol use, drug use, tobacco use, obesity, dyslipidemia, pneumonia complication.

When comparing race/ethnic and sex-specific in-hospital mortality by hospital categories, there were no differences in in-hospital mortality between White women and White men regardless of hospital type (**Table 4**). In all hospitals, non-White men and women had lower adjusted mortality than white men (aOR 0.89, 0.84-0.94, p<0.001, aOR 0.89, 0.84-0.94, p<0.001, respectively; **Table 4**).

**Table 4.** Adjusted comparisons of in-hospital mortality odds across race/ethnicity and sex by hospital category, 2006-2017.

| Proportion of non-White patients served by hospital | Race/ethnicity and sex | In-hospital mortality adjusted OR<sup>†</sup> (95% CI) | p-value |
| --- | --- | --- | --- |
| ≤25% | White men | Ref | Ref |
|  | Non-White men | 0.86 (0.77, 0.97) | 0.012 |
|  | White women | 1.02 (0.97, 1.07) | 0.409 |
|  | Non-White women | 0.78 (0.70, 0.87) | <0.001 |
| 25-50% | White men | Ref | Ref |
|  | Non-White men | 0.88 (0.79, 1.00) | 0.010 |
|  | White women | 1.01 (0.93, 1.09) | 0.829 |
|  | Non-White women | 0.84 (0.76, 0.92) | <0.001 |
| >50% | White men | Ref | Ref |
|  | Non-White men | 0.82 (0.74, 0.92) | 0.001 |
|  | White women | 0.98 (0.87, 1.11) | 0.754 |
|  | Non-White women | 0.90 (0.81, 1.00) | 0.043 |
| All Hospitals | White men | Ref | Ref |
|  | Non-White men | 0.89 (0.84, 0.94) | <0.001 |
|  | White women | 1.02 (0.98, 1.06) | 0.347 |
|  | Non-White women | 0.89 (0.84, 0.94) | <0.001 |

<sup>†</sup>Adjusted for age, sex, insurance type, income by zip code, hospital region, hospital location/teaching status, hospital ownership, hospital bed size, APRDRG Severity, APRDRG mortality risk, Charlson Comorbidity Index, DNR, palliative care, alcohol use, drug use, tobacco use, obesity, dyslipidemia, pneumonia complication.

Finally, when the adjusted odds of in-hospital mortality were compared between hospital strata, patients admitted to hospitals predominantly serving non-White patients had higher odds of in-hospital mortality compared to those admitted to hospitals predominantly serving White patients (aOR 1.08, 1.01-1.15, p=0.021) (**Table 5**). This difference was driven by the Hispanic and API patients, who had 45% and 36% higher odds of mortality, respectively, if they were admitted to a hospital predominantly serving non-White patients compared to being admitted to a hospital predominantly serving White patients (aOR 1.45, 1.18-1.78, p<0.001, aOR 1.36, 1.00-1.86, p=0.050, respectively).

**Table 5.** Adjusted comparisons of in-hospital mortality according to hospital type within each race/ethnicity.

| Race/ethnicity | % non-White patients served by hospital | In-hospital mortality adjusted OR<sup>†</sup> (95% CI) | p-value |
| --- | --- | --- | --- |
| All | ≤25% | Ref | Ref |
|  | 25-50% | 1.02 (0.97, 1.08) | 0.417 |
|  | >50% | 1.08 (1.01, 1.15) | 0.021 |
| White | ≤25% | Ref | Ref |
|  | 25-50% | 1.03 (0.97, 1.09) | 0.381 |
|  | >50% | 1.07 (0.99, 1.16) | 0.110 |
| Black | ≤25% | Ref | Ref |
|  | 25-50% | 1.05 (0.91, 1.20) | 0.517 |
|  | >50% | 1.00 (0.87, 1.15) | 0.972 |
| Hispanic | ≤25% | Ref | Ref |
|  | 25-50% | 1.18 (0.94, 1.48) | 0.146 |
|  | >50% | 1.45 (1.18, 1.78) | <0.001 |
| Asian/Pacific Islander | ≤25% | Ref | Ref |
|  | 25-50% | 1.10 (0.81, 1.50) | 0.541 |
|  | >50% | 1.36 (1.00, 1.86) | 0.050 |

<sup>†</sup> Adjusted for age, sex, insurance type, income by zip code, hospital region, hospital location/teaching status, hospital ownership, hospital bed size, APRDRG Severity, APRDRG mortality risk, Charlson Comorbidity Index, DNR, palliative care, alcohol use, drug use, tobacco use, obesity, dyslipidemia, pneumonia complication.

## Discussion

From 2006 to 2017, in-hospital mortality odds after AIS declined steadily by 67%. Although in-hospital mortality declined for all race/ethnic subgroups, the decline was less pronounced for Black individuals. Contrary to our initial hypothesis, Black and Hispanic AIS patients had lower odds of in-hospital mortality than their White counterparts, and the difference was primarily driven by the elderly and very elderly age groups. The survival advantage of the Black and Hispanic individuals was most pronounced in hospitals predominantly serving White patients and least in hospitals mainly serving non-White patients. AIS patients of all race/ethnicities had higher mortality when they were admitted to hospitals serving predominantly non-White individuals. When stratified by race/ethnicity, Hispanic and API individuals had higher mortality rates if they were admitted to hospitals serving predominantly non-White individuals.

Contrary to our findings, another NIS of a shorter, more recent time frame showed that from 2016 to 2020 in-hospital mortality among AIS patients increased slightly, and this increase was found predominantly in those who were Hispanic, had COVID-19, and low income^29^. Past epidemiologic studies demonstrated an improvement in stroke mortality in Black individuals overall^2,4,5^ and in women^2,6^ from 1996 to 2015.

Our findings of higher in-hospital mortality in hospitals serving predominantly non-White individuals, suggests that system-level effects are at play on patients regardless of their demographics. This is consistent with previous studies in patients with stroke, coronary artery bypass graft, myocardial infarction, and trauma, suggesting that institutions predominantly serving Black and Hispanic individuals yielded poorer outcomes.^12–14,17^

Our study demonstrated higher in-hospital mortality for all race/ethnicities in hospitals predominantly serving non-White individuals; this could be due to decisions regarding withdrawal of care. We used DNR and palliative care orders as surrogate markers of withdrawal of care. Prior studies have shown differences by race/ethnicity in use of DNR and palliative care orders in stroke patients. An NIS study of individuals with hemorrhagic strokes showed that White patients are more likely to receive DNR or palliative orders^30^. A study in 2006-2014 showed that DNR orders in AIS were less likely to be given to Mexican Americans and Black individuals within 24 hours, but no difference was found afterwards^31^. Another study reported lower utilization of palliative care in 2007-2011 in non-White minority patients in all hospital strata by the proportion of the non-White patients served, as well as in all race/ethnicities in hospitals predominantly serving the non-White patients^32^.

The temporal decline in in-hospital mortality may reflect improvement in systems of care, including the proliferation of primary and comprehensive stroke centers^33,34^, increasing use of intravenous thrombolysis^35,36^ and endovascular therapy^37,38^, and improved pre-hospital care^39^. The less pronounced temporal decline in in-hospital mortality for Black individuals could be partially explained by racial inequities in prehospital care^40^ and treatment with intravenous thrombolysis^17^. The lower in-hospital mortality for Black and Hispanic individuals could reflect disparate mechanisms of stroke. Black and Hispanic individuals have higher proportion of small vessel disease while White individuals have higher proportion of atrial fibrillation^41^ and cardioembolism^42^, which tend to result in more severe strokes with a higher chance of death compared to small vessel strokes^43^.

This study is limited by the cross-sectional design and use of administrative data, which are prone to coding errors. Additionally, the time frame in this study included the transition from ICD-9 to ICD-10 coding eras, potentially resulting in discrepancies in patient inclusion. However, ICD-9 and ICD-10 codes have been shown to detect cerebrovascular disease with a sensitivity of ≥82% and specificity of ≥95%^44^. The sampling structure of the NIS changed in 2012 to sample 20% of all discharges, rather than 20% of all hospitals.^45^ This could have led to inaccuracies in the categorization of the hospitals by the proportion of non-White patients. The NIS does not collect measures of stroke-specific disease severity, such as National Institutes of Health Stroke Scale, which has been shown to predict in-hospital mortality^46^. Despite these limitations, the study strengths include the nationwide representative nature of NIS and adjustment for key clinical factors that may impact in-hospital mortality, including DNR status and palliative care.

## Data Availability

The raw data supporting the conclusions of this article will be made available by the authors, without undue reservation.

## Acknowledgments

This study was funded by the James and Dorothy Williams Stroke Scholarship.

## References

1. Virani SS, Alonso A, Benjamin EJ, Bittencourt MS, Callaway CW, Carson AP, Chamberlain AM, Chang AR, Cheng S, Delling FN, et al. Heart Disease and Stroke Statistics-2020 Update: A Report From the American Heart Association. Circulation. 2020;141:e139–e596. doi: 10.1161/CIR.0000000000000757

2. Towfighi A, Ovbiagele B, Saver JL. Therapeutic milestone: stroke declines from the second to the third leading organ- and disease-specific cause of death in the United States. Stroke. 2010;41:499–503. doi: 10.1161/STROKEAHA.109.571828

3. Towfighi A, Saver JL. Stroke declines from third to fourth leading cause of death in the United States: historical perspective and challenges ahead. Stroke. 2011;42:2351–2355. doi: 10.1161/STROKEAHA.111.621904

4. Lackland DT, Roccella EJ, Deutsch AF, Fornage M, George MG, Howard G, Kissela BM, Kittner SJ, Lichtman JH, Lisabeth LD, et al. Factors influencing the decline in stroke mortality: a statement from the American Heart Association/American Stroke Association. Stroke. 2014;45:315–353. doi: 10.1161/01.str.0000437068.30550.cf

5. Carnethon MR, Pu J, Howard G, Albert MA, Anderson CAM, Bertoni AG, Mujahid MS, Palaniappan L, Taylor HA, Jr., Willis M, et al. Cardiovascular Health in African Americans: A Scientific Statement From the American Heart Association. Circulation. 2017;136:e393–e423. doi: 10.1161/CIR.0000000000000534

6. Arnao V, Acciarresi M, Cittadini E, Caso V. Stroke incidence, prevalence and mortality in women worldwide. Int J Stroke. 2016;11:287–301. doi: 10.1177/1747493016632245

7. Bach PB, Pham HH, Schrag D, Tate RC, Hargraves JL. Primary care physicians who treat blacks and whites. N Engl J Med. 2004;351:575–584. doi: 10.1056/NEJMsa040609

8. Groeneveld PW, Laufer SB, Garber AM. Technology diffusion, hospital variation, and racial disparities among elderly Medicare beneficiaries: 1989-2000. Med Care. 2005;43:320–329. doi: 10.1097/01.mlr.0000156849.15166.ec

9. Rothenberg BM, Pearson T, Zwanziger J, Mukamel D. Explaining disparities in access to high-quality cardiac surgeons. Ann Thorac Surg. 2004;78:18–24; discussion 24-15. doi: 10.1016/j.athoracsur.2004.01.021

10. Skinner J, Chandra A, Staiger D, Lee J, McClellan M. Mortality after acute myocardial infarction in hospitals that disproportionately treat black patients. Circulation. 2005;112:2634–2641. doi: 10.1161/CIRCULATIONAHA.105.543231

11. Bradley EH, Herrin J, Wang Y, McNamara RL, Webster TR, Magid DJ, Blaney M, Peterson ED, Canto JG, Pollack CV, Jr., et al. Racial and ethnic differences in time to acute reperfusion therapy for patients hospitalized with myocardial infarction. JAMA. 2004;292:1563–1572. doi: 10.1001/jama.292.13.1563

12. Konety SH, Vaughan Sarrazin MS, Rosenthal GE. Patient and hospital differences underlying racial variation in outcomes after coronary artery bypass graft surgery. Circulation. 2005;111:1210–1216. doi: 10.1161/01.CIR.0000157728.49918.9F

13. Barnato AE, Lucas FL, Staiger D, Wennberg DE, Chandra A. Hospital-level racial disparities in acute myocardial infarction treatment and outcomes. Med Care. 2005;43:308–319. doi: 10.1097/01.mlr.0000156848.62086.06

14. Haider AH, Ong’uti S, Efron DT, Oyetunji TA, Crandall ML, Scott VK, Haut ER, Schneider EB, Powe NR, Cooper LA, et al. Association between hospitals caring for a disproportionately high percentage of minority trauma patients and increased mortality: a nationwide analysis of 434 hospitals. Arch Surg. 2012;147:63–70. doi: 10.1001/archsurg.2011.254

15. Baicker K, Chandra A, Skinner JS, Wennberg JE. Who you are and where you live: how race and geography affect the treatment of medicare beneficiaries. Health Aff (Millwood). 2004;Suppl Variation:VAR33–44. doi: 10.1377/hlthaff.var.33

16. Epstein AM. Health care in America--still too separate, not yet equal. N Engl J Med. 2004;351:603–605. doi: 10.1056/NEJMe048181

17. Faigle R, Urrutia VC, Cooper LA, Gottesman RF. Individual and System Contributions to Race and Sex Disparities in Thrombolysis Use for Stroke Patients in the United States. Stroke. 2017;48:990–997. doi: 10.1161/STROKEAHA.116.015056

18. HCUP National Inpatient Sample (NIS). Healthcare Cost and Utilization Project (HCUP). Agency for Healthcare Research and Quality. www.hcup-us.ahrq.gov/nisoverview.jsp. 2006-2017. Accessed 03/01.

19. Quan H, Sundararajan V, Halfon P, Fong A, Burnand B, Luthi JC, Saunders LD, Beck CA, Feasby TE, Ghali WA. Coding algorithms for defining comorbidities in ICD-9-CM and ICD-10 administrative data. Med Care. 2005;43:1130–1139. doi: 10.1097/01.mlr.0000182534.19832.83

20. Deyo RA, Cherkin DC, Ciol MA. Adapting a clinical comorbidity index for use with ICD-9-CM administrative databases. J Clin Epidemiol. 1992;45:613–619. doi: 10.1016/0895-4356(92)90133-8

21. Goldstein LB, Samsa GP, Matchar DB, Horner RD. Charlson Index comorbidity adjustment for ischemic stroke outcome studies. Stroke. 2004;35:1941–1945. doi: 10.1161/01.STR.0000135225.80898.1c

22. McCormick PJ, Lin HM, Deiner SG, Levin MA. Validation of the All Patient Refined Diagnosis Related Group (APR-DRG) Risk of Mortality and Severity of Illness Modifiers as a Measure of Perioperative Risk. J Med Syst. 2018;42:81. doi: 10.1007/s10916-018-0936-3

23. Xian Y, Holloway RG, Pan W, Peterson ED. Challenges in assessing hospital-level stroke mortality as a quality measure: comparison of ischemic, intracerebral hemorrhage, and total stroke mortality rates. Stroke. 2012;43:1687–1690. doi: 10.1161/STROKEAHA.111.648600

24. Bar B, Hemphill JC, 3rd. Charlson comorbidity index adjustment in intracerebral hemorrhage. Stroke. 2011;42:2944–2946. doi: 10.1161/STROKEAHA.111.617639

25. Cassel JB, Jones AB, Meier DE, Smith TJ, Spragens LH, Weissman D. Hospital mortality rates: how is palliative care taken into account? J Pain Symptom Manage. 2010;40:914–925. doi: 10.1016/j.jpainsymman.2010.07.005

26. Qureshi AI, Adil MM, Suri MF. Rate of utilization and determinants of withdrawal of care in acute ischemic stroke treated with thrombolytics in USA. Med Care. 2013;51:1094–1100. doi: 10.1097/MLR.0b013e3182a95db4

27. Stubbs JM, Assareh H, Achat HM, Greenaway S, Muruganantham P. Verification of administrative data to measure palliative care at terminal hospital stays. Health Inf Manag. 2020:1833358320968572. doi: 10.1177/1833358320968572

28. von Elm E, Altman DG, Egger M, Pocock SJ, Gotzsche PC, Vandenbroucke JP, Initiative S. The Strengthening the Reporting of Observational Studies in Epidemiology (STROBE) statement: guidelines for reporting observational studies. Ann Intern Med. 2007;147:573–577. doi: 10.7326/0003-4819-147-8-200710160-00010

29. de Havenon A, Zhou LW, Yaghi S, Frontera JA, Sheth KN. Effect of COVID-19 on Acute Ischemic Stroke Severity and Mortality in 2020: Results From the 2020 National Inpatient Sample. Stroke. 2023;54:e194–e198. doi: 10.1161/STROKEAHA.122.041929

30. Cruz-Flores S, Rodriguez GJ, Chaudhry MRA, Qureshi IA, Qureshi MA, Piriyawat P, Vellipuram AR, Khatri R, Kassar D, Maud A. Racial/ethnic disparities in hospital utilization in intracerebral hemorrhage. Int J Stroke. 2019;14:686–695. doi: 10.1177/1747493019835335

31. Bailoor K, Shafie-Khorassani F, Lank RJ, Case E, Garcia NM, Lisabeth LD, Sanchez BN, Kim S, Morgenstern LB, Zahuranec DB. Time Trends in Race-Ethnic Differences in Do-Not-Resuscitate Orders After Stroke. Stroke. 2019;50:1641–1647. doi: 10.1161/STROKEAHA.118.024460

32. Faigle R, Ziai WC, Urrutia VC, Cooper LA, Gottesman RF. Racial Differences in Palliative Care Use After Stroke in Majority-White, Minority-Serving, and Racially Integrated U.S. Hospitals. Crit Care Med. 2017;45:2046–2054. doi: 10.1097/CCM.0000000000002762

33. Uchino K, Man S, Schold JD, Katzan IL. Stroke Legislation Impacts Distribution of Certified Stroke Centers in the United States. Stroke. 2015;46:1903–1908. doi: 10.1161/STROKEAHA.114.008007

34. Schieb LJ, Casper ML, George MG. Mapping Primary and Comprehensive Stroke Centers by Certification Organization. Circ Cardiovasc Qual Outcomes. 2015;8:S193–194. doi: 10.1161/CIRCOUTCOMES.115.002082

35. Schwamm LH, Ali SF, Reeves MJ, Smith EE, Saver JL, Messe S, Bhatt DL, Grau-Sepulveda MV, Peterson ED, Fonarow GC. Temporal trends in patient characteristics and treatment with intravenous thrombolysis among acute ischemic stroke patients at Get With The Guidelines-Stroke hospitals. Circ Cardiovasc Qual Outcomes. 2013;6:543–549. doi: 10.1161/CIRCOUTCOMES.111.000303

36. George BP, Asemota AO, Dorsey ER, Haider AH, Smart BJ, Urrutia VC, Schneider EB. United States trends in thrombolysis for older adults with acute ischemic stroke. Clin Neurol Neurosurg. 2015;139:16–23. doi: 10.1016/j.clineuro.2015.08.031

37. Shah S, Xian Y, Sheng S, Zachrison KS, Saver JL, Sheth KN, Fonarow GC, Schwamm LH, Smith EE. Use, Temporal Trends, and Outcomes of Endovascular Therapy After Interhospital Transfer in the United States. Circulation. 2019;139:1568–1577. doi: 10.1161/CIRCULATIONAHA.118.036509

38. To CY, Rajamand S, Mehra R, Falatko S, Badr Y, Richards B, Qahwash O, Fessler RD. Outcome of mechanical thrombectomy in the very elderly for the treatment of acute ischemic stroke: the real world experience. Acta Radiol Open. 2015;4:2058460115599423. doi: 10.1177/2058460115599423

39. Demaerschalk BM, Scharf EL, Cloft H, Barrett KM, Sands KA, Miller DA, Meschia JF. Contemporary Management of Acute Ischemic Stroke Across the Continuum: From TeleStroke to Intra-Arterial Management. Mayo Clin Proc. 2020;95:1512–1529. doi: 10.1016/j.mayocp.2020.04.002

40. Bhattacharya P, Mada F, Salowich-Palm L, Hinton S, Millis S, Watson SR, Chaturvedi S, Rajamani K. Are racial disparities in stroke care still prevalent in certified stroke centers? J Stroke Cerebrovasc Dis. 2013;22:383–388. doi: 10.1016/j.jstrokecerebrovasdis.2011.09.018

41. Shen AY, Contreras R, Sobnosky S, Shah AI, Ichiuji AM, Jorgensen MB, Brar SS, Chen W. Racial/ethnic differences in the prevalence of atrial fibrillation among older adults--a cross-sectional study. J Natl Med Assoc. 2010;102:906–913. doi: 10.1016/s0027-9684(15)30709-4

42. Gutierrez J, Koch S, Dong C, Casanova T, Modir R, Katsnelson M, Ortiz GA, Sacco RL, Romano JG, Rundek T. Racial and ethnic disparities in stroke subtypes: a multiethnic sample of patients with stroke. Neurol Sci. 2014;35:577–582. doi: 10.1007/s10072-013-1561-z

43. Arboix A, Alio J. Cardioembolic stroke: clinical features, specific cardiac disorders and prognosis. Curr Cardiol Rev. 2010;6:150–161. doi: 10.2174/157340310791658730

44. McCormick N, Bhole V, Lacaille D, Avina-Zubieta JA. Validity of Diagnostic Codes for Acute Stroke in Administrative Databases: A Systematic Review. PLoS One. 2015;10:e0135834. doi: 10.1371/journal.pone.0135834

45. Khera R, Angraal S, Couch T, Welsh JW, Nallamothu BK, Girotra S, Chan PS, Krumholz HM. Adherence to Methodological Standards in Research Using the National Inpatient Sample. JAMA. 2017;318:2011–2018. doi: 10.1001/jama.2017.17653

46. Kortazar-Zubizarreta I, Pinedo-Brochado A, Azkune-Calle I, Aguirre-Larracoechea U, Gomez-Beldarrain M, Garcia-Monco JC. Predictors of in-hospital mortality after ischemic stroke: A prospective, single-center study. Health Sci Rep. 2019;2:e110. doi: 10.1002/hsr2.110

